# “I can be a source of motivation”: Perspectives from stakeholders of the I’mPossible fellowship, a peer-led differentiated service delivery model for adolescents with perinatally acquired HIV in India

**DOI:** 10.1101/2025.03.11.25323808

**Authors:** Siddha Sannigrahi, Michael Babu Raj, Babu Seenappa, Ashley A. Sharma, Suhas Reddy, Esha Nobbay, Aastha Kant, Satish Kumar SK, Baldeep K. Dhailwal, Lakshmi Ganapathi, Anita Shet

## Abstract

Youth living with HIV (YLHIV) face multidimensional challenges, including stigma, mental health struggles, and socioeconomic instability, which are further magnified among adolescents and young adults with perinatally acquired HIV (APHIV). Peer-led differentiated service delivery (DSD) models providing tailored support for this population have demonstrated improved outcomes, though their adaptation and implementation in India remain underexplored. We examined multi-stakeholder perceptions within a peer-support DSD intervention in India, the I’mPossible Fellowship, designed to address the health, educational, and livelihood needs of APHIV. Between May and December 2023, we enrolled three stakeholder groups involved in the I’mPossible fellowship intervention: (1) intervention deliverers (APHIV “fellows”), (2) facilitators (“supervisors” of APHIV), and (3) beneficiaries (APHIV “peers”). To examine fellows’ roles and growth, we conducted in-depth interviews with fellows (n=8; 75% female; mean age: 22.5 years) and supervisors (n=7). Aiming to explore fellow-peer interactions and perceived program impact, we conducted three focus group discussions with purposefully sampled peers (n=18; 66.7% male; mean age: 16.7 years). Data collected were audio recorded, transcribed, and translated from Kannada to English for coding and analysis. Thematic deductive analysis was combined with data triangulation across participant groups to synthesize findings. Five key themes emerged, highlighting the layered influences of the I’mPossible Fellowship. First, mentorship was an important theme that provided informational and emotional support for peers; second, peer influence arising from peer-to-peer interactions contributed to a sense of trust and affirmation. Third, personal growth experienced by fellows stimulated their motivation to fulfil their mentorship roles effectively. Fourth, complex systemic challenges, such as stigma and discrimination, hindered educational and employment advancement of APHIV. Fifth, sustainability, through robust post-fellowship systems and continued mentoring support, was emphasized by supervisors and fellows as crucial for supporting APHIV in transitioning to independent living. This study highlights the pivotal role of fellows and the bi-directional power of peer mentorships in addressing the multilevel factors that enhance outcomes for APHIV. By providing knowledge and empathy to their peers and serving as credible role models with lived experience of HIV, fellows within the I’mPossible fellowship exemplify a successful DSD model incorporating the three essential attributes of peer support: informational, emotional, and affirmative support. While these findings underscore the importance of integrating peer-led interventions into HIV care frameworks to support youth with HIV, this also reframes youth as active agents of change, recognizing their capacity for empowerment and meaningful societal contribution rather than passive recipients of care.

## Introduction

An estimated 3.1 million adolescents and young adults ages 15 to 24 years old were living with human immunodeficiency virus (HIV) globally in 2023 (1). With the advancement and increased availability of antiretroviral treatment (ART) options, a more significant number of youth living with HIV (YLHIV) are surviving into adulthood (2,3). This demographic confronts profound psychosocial and health-related challenges, such as the compounded stress of managing a stigmatizing, chronic illness, alongside experiences of parental loss, financial instability, and social discrimination, that significantly exacerbate their vulnerability (4–8). Transitioning into adult healthcare systems frequently presents logistical and emotional hurdles that can further complicate long-term care engagement (7,9,10). These adversities manifest in reduced adherence to ART and poor viral suppression rates, leading to higher rates of treatment failure, morbidity, and mortality compared to other age groups (11–14).

HIV care models have sought to address these multifaceted challenges among YLHIV using differentiated service delivery (DSD) models in HIV care (14). These models adapt the provision of services—such as medication dispensing, counseling, and follow-up care—to better align with their unique lifestyles, preferences, and needs, offering a more person-centered approach to healthcare. Peer support has emerged as an effective strategy to tailor services to individual needs, as it leverages shared lived experiences to foster trust, reduce isolation, and provide both practical and emotional guidance (12,15–17). This support can be leveraged through community-based DSD models, as opposed to facility-based ones, and is more advantageous in improving accessibility by improving barriers such as travel costs and rigid scheduling (18–20). This positions YLHIV not merely as disadvantaged recipients of care but as individuals with agency capable of being empowered to contribute meaningfully as social assets within their communities.

Research from sub-Saharan Africa highlights the potential of peer support interventions to improve viral suppression and ART adherence among YLHIV. However, there are critical gaps in the existing literature (19,21). First, while existing peer support models demonstrate progress in improving health outcomes among YLHIV, their broader psychosocial, educational, and livelihood needs extend beyond medical care and are often overlooked in research and programmatic implementation. Second, in many HIV peer support programs, a recurring challenge is the limited focus on “caring for the carer,” referring to the emotional and practical needs of the peer supporters who deliver care and guidance to their communities (22). While these carers provide substantial support to YLHIV, programs often neglect strategies to sustain their well-being, leaving them vulnerable to emotional exhaustion. Third, studies on peer support are centered mainly around experiences in the Sub-Saharan Africa region, while the YLHIV population in India remains under-recognized. India has the largest population of YLHIV in Asia, with national estimates from 2023 indicating 163,000 adolescents and young adults aged 15-24 years are living with HIV (2,23,24). In particular, a large proportion are adolescents and young adults with perinatally acquired HIV (APHIV), a distinct subgroup of youth with HIV diagnosed in early childhood.

Our study addresses these three gaps–support beyond health needs, focus on the intervention deliverer, geographic gap in India–by studying the I’mPossible fellowship program, a peer-led intervention for APHIV in India. In this study, we examine experiences of fellows providing peer support and triangulate findings with qualitative data obtained from beneficiaries of this support and from stakeholders who function as informal mentors to fellows. Through this well-rounded qualitative exploration, this research aims to reveal the practical realities of implementing peer-led DSD models and the potential of these models to inform improved HIV care in India and similar settings.

## Methods

### Study Setting

This study was conducted in six districts across the south Indian states of Karnataka and Tamil Nadu (Bangalore, Belgaum, Krishnagiri, Gulbarga, Kolar, Vijaypura, and Raichur) between 3 May 2023 to 24 November 2023. Procedures were executed in collaboration with local partners, non-profit organizations, families of APHIV, and childcare institutions (CCIs) that offer residential support and care for APHIV in these regions.

### Overview of the I’mPossible fellowship

The I’mPossible fellowship, launched in 2021, is a peer-support DSD model designed and adapted to support APHIV in the southern Indian states of Karnataka and Tamil Nadu. Using a hybrid delivery model comprised of one-on-one and group sessions conducted over a 48-week period, children, adolescents, and young adults who are living with HIV, referred to as “peers”, are provided with tailored health, educational, and livelihood support. The program is implemented by trained youth aged 18 to 24, known as “fellows”, who are living with HIV. Fellows are recruited based on their demonstrated motivation and leadership potential. They then engage in a month-long in-house training at a partner institution based on sessions adapted from the International Center for AIDS Care and Treatment Programs curriculum (25). Following the training, fellows are assigned approximately 25 peers residing either in CCIs or within the community in family-based care. Based on their location, each fellow serves as a peer mentor and works under the guidance of an experienced supervisor who provides oversight and support. Fellows are supported through stipends and encouraged to maintain their health and pursue higher education or vocational training. This dual-focus approach ensures the program’s sustainability by prioritizing fellows’ well-being and professional development while enabling them to act as agents of change within their communities.

### I’mPossible fellowship and the social-ecological model of health

The I’mPossible fellowship intervention is grounded in the socio-ecological model (SEM) of health, which serves as a person- and context-centered intervention framework (26). This framework addresses the individual while emphasizing the multiple spheres of influence (interpersonal, organizational, community, and public policy) on health and well-being (26). The fellowship focuses on three levels - individual, interpersonal, and organizational - and integrates stakeholders within each domain (Figure 1). Peers represent the personal level, reflecting their unique knowledge, attitudes, and skills in self-managing HIV. Fellows embody the interpersonal level, highlighting the interactive dynamics and support they provide to peers. Supervisors, who often fulfill the role of guardians for peers and as informal mentors to fellows, operate within organizational and supervisory structures such as HIV clinical care centers, religious communities, and non-profit organizations representing the interpersonal and organizational levels of influence.

**FIGURE 1.**
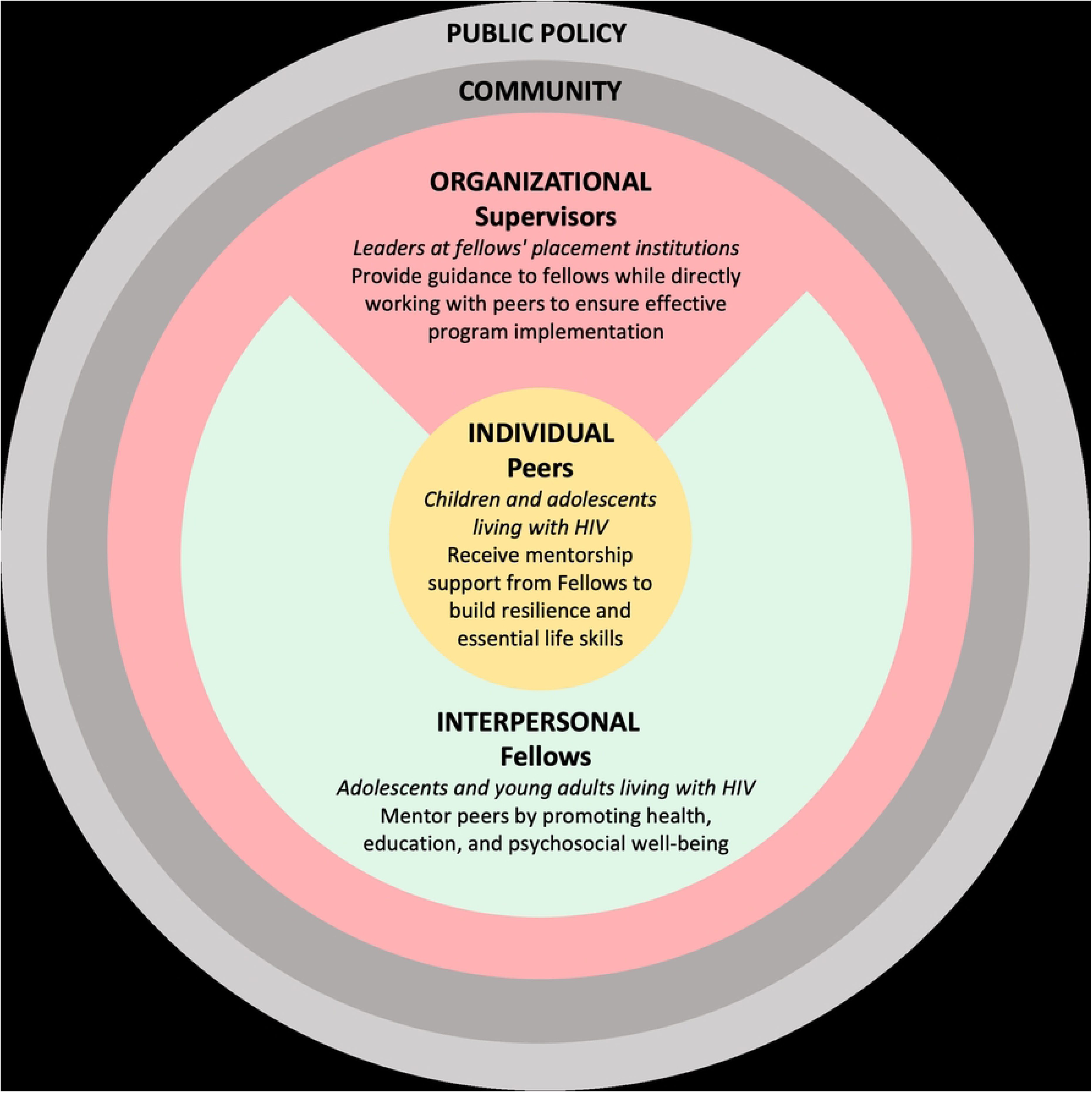
Study participants’ perspectives presented through the lens of the socio-ecological model framework.

### Study participants

This study presents qualitative accounts from three key stakeholder groups involved in the I’mPossible fellowship—fellows, peers, and supervisors—each representing a distinct perspective within the socio-ecological model (Figure 1). In this study, the term “YLHIV”’ is used to refer to all youth living with HIV, whereas “APHIV” denotes the subset of YLHIV who acquired HIV perinatally, defined as those diagnosed with HIV prior to the age of 10 years. All Fellows and peer participants in this study are APHIV by this definition.

Eligible fellows were those from the first cohort of the I’mPossible fellowship, and peers were APHIV aged <18 years who maintained regular contact with at least one fellow from the I’mPossible intervention for a minimum of three months. Supervisors were purposively selected from faculty or staff members at CCIs, non-governmental organization managers, and healthcare workers who directly supervised the fellow for at least three months.

### Ethics approval and consent to participate

The study received approval from the Institutional Review Boards of the YR Gaitonde Centre for AIDS Research and Education in Chennai, Tamil Nadu (#YRG375) and the Johns Hopkins Bloomberg School of Public Health in the United States (#IRB00023077). Informed consent for this study was obtained orally due to the combination of in-person and virtual data collection, which made obtaining written consent logistically challenging, as virtual participants often lacked access to printers or scanners to provide signed consent forms. Oral consent, assent, and parental permission were obtained through researcher-signed documentation, which confirmed that informed consent was provided and that participants voluntarily agreed to participate in the study. Adult participants provided oral informed consent for their participation. For participants <18 years, oral parental permission was obtained from their parents or legal guardians, and oral assent from the minor participants themselves. Institutional Review Boards approved the use of oral consent, assent, and parental permission ensuring that participants received a clear explanation of the study’s purpose, procedures, risks, and benefits.

### Data Collection

To support the multi-perspective qualitative research on the impact of the I’mPossible intervention, we first conducted individual in-depth interviews (IDIs) with fellows (n=8; 75% female; mean age: 22.5 years) and supervisors (n=7) to (1) explore fellows’ motivations, leadership development, and overall experiences within the intervention, and (2) examine the roles and contributions of fellows, their perception of the influence of the intervention on peers, specifically assessing observable benefits or changes in the fellows and peers. Interviews were conducted in English by a trained interviewer (SS) either over the phone or at a location preferred by the participants. The interviews were continued until saturation was reached and lasted approximately 1–2 hours per participant. All fellows and supervisors were proficient in English. Next, we conducted three focus group discussions (FGDs) with peers (n=18; 66.7% male; mean age: 16.7 years) to characterize their interactions with fellows, the program’s impact on their quality of life, and the challenges of living with HIV. FGDs were facilitated by trained India-based researchers (MBR, EN) in Kannada and English. Interview guides for IDIs and FGDs were semi-structured and iteratively developed with input from program staff and youth investigators from the community (BS, SR). All sessions were audio-recorded with participants’ written informed consent. The research team (SR, SS, EN) transcribed the audio recordings, and translation services were facilitated by a local translator based in Karnataka.

### Data Analysis

The research team coded and analyzed the IDI and FGD transcripts using Dedoose 9.0 software. Two complementary methods of analysis were employed to assess the multiple-perspective qualitative data. First, conducting a thematic analysis, following the methodology outlined by Braun & Clarke (2006) (27). Independent coders (SS, AAS, AK) carried out the analysis in six iterative steps: they familiarized themselves with the data, generated initial codes, searched for themes, reviewed themes, defined and named themes, and produced the final report for each data collection method. Second, they applied the analytical steps for triangulating qualitative multiple-perspective interviews, as described by Vogl, Schmidt, and Zartler (2019), was used (28). This approach further examined the coded data generated through the thematic analysis across different participant groups (fellows, peers, and supervisors) to identify patterns, similarities, and contrasts among their perspectives (28). To ensure that participant experiences were accurately captured, the researchers conducted member checking, also known as participant validation, for all data collection activities. Additionally, researchers engaged in reflexivity throughout the research process through qualitative memo writing and regular debriefing sessions to discuss the potential influence of their positionality on data collection and analysis.

## Results

Fellows, supervisors, and peers provided converging yet distinct perspectives on the core themes of the I’mPossible fellowship intervention. While all three groups recognized the importance of mentorship and emotional support within the fellowship, their perspectives highlighted specific insights regarding the intervention and generated recommendations on how the fellowship can better support the APHIV community. Their collective experiences revealed the fellowship’s multi-layered effects through five core themes: (1) mentorship, (2) peer influence, (3) personal growth, (4) complex challenges, and (5) sustainability (Tables 1 and 2).

**Table 1.**
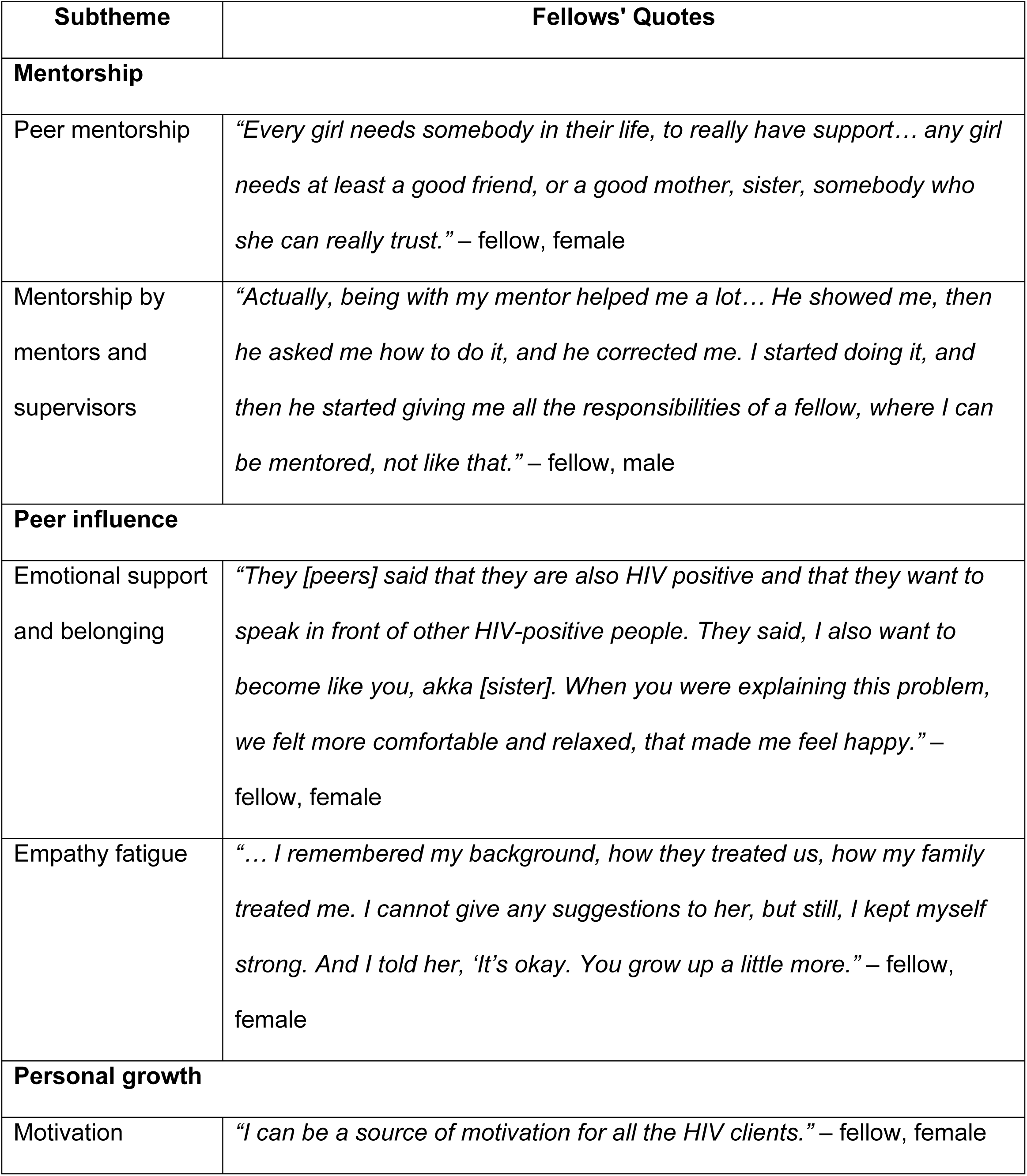
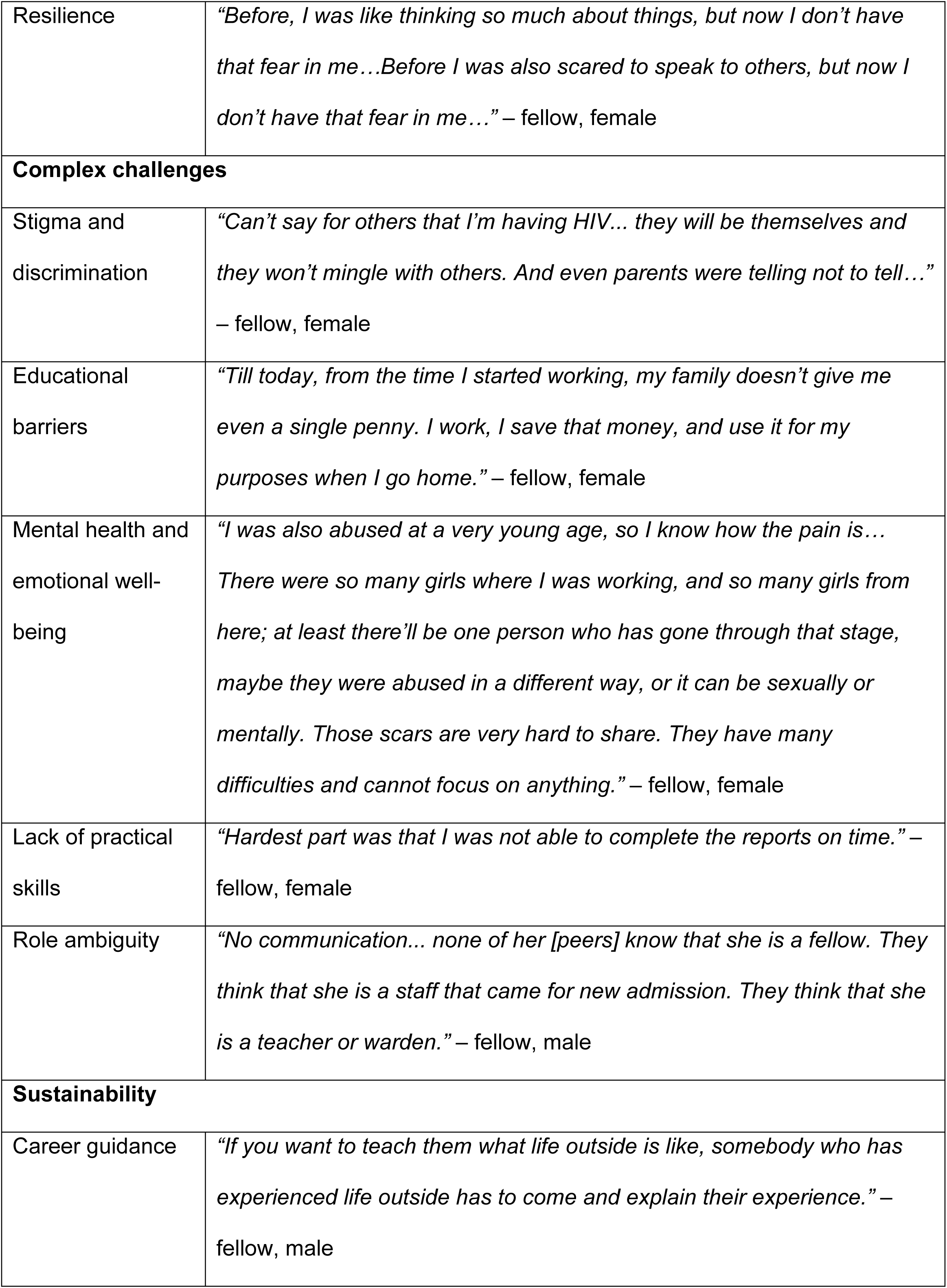
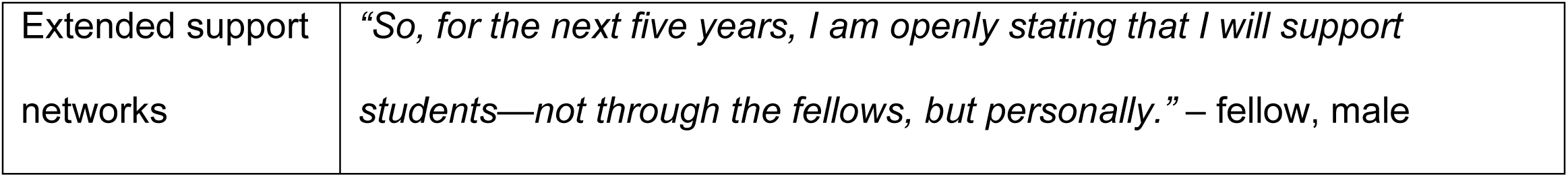
Main themes and subthemes from fellows’ perspectives with exemplary quotes.

**Table 2.** Themes and exemplary quotes from supervisors and peers, organized by sample.

| Theme | Finding | Supervisor Quote | Peer Quote |
| --- | --- | --- | --- |
| **Mentorship** | Mentorship fosters informational, emotional and academic support. | *“... she [fellow] herself was giving her own examples and telling them [peers]... I have been taking these tablets for so many years.” -*<br>– supervisor 1 | *“She [fellow] was helping us in our studies.” –* peer, male |
| **Peer influence** | Shared lived experiences build trust and engagement. | *“...these people [fellows] are always available, and at the same time, they are ready to understand their own person [peer] who is suffering from the same difficulties.” –*<br>supervisor 3 | *“With [HIV] positive people [such as fellows], we can speak much more freely.” –*<br>peer, female |
| **Personal growth** | Fellows develop leadership and resilience through | *“It was a real learning for her, like, see, even our fellows, like, they were just like the children previously in the institutions, and now* | *“[ If we were to become fellows] We will get confidence by teaching others and help others by* |
|  | carrying out their responsibilities. | *they themselves are monitoring this.” – supervisor 1* | *telling them to take tablets.” – peer, male* |
| **Complex challenges** | Stigma and educational barriers create uncertainty about the future, which may impede outcomes. | *Stigma and negative ideas are there, and nobody is educating them. Nobody is bringing awareness. Awareness is there for the ones who are positive, but not for the family members... – supervisor 2* | *“First we were studying in another school and once they came to know about our status, then we were put in another school.” – peer, male* |
| **Sustainability** | Strengthened community connections support long-term success. | *“I feel we should be like...somehow we have to gather them once in a while or..., in some way we have to just give them...mentoring and helping them to stay connected.” – supervisor 5* | *“Now we are in [CCI]. Here we get support but after we go out from [CCI] where will we get support from?” – peer, male* |

### Mentorship

Mentorship, built foundationally on trust, was central to the fellowship’s impact. Fellows stepped into dual roles as both teachers and learners, providing emotional and practical guidance while also bidirectionally benefiting from mentorship by supervisors and program staff. Peers valued this dynamic and regarded fellows as mentors, frequently seeking their guidance not only for medical and HIV-related issues, but also for educational and emotional support.

More than just sources of information, fellows created safe spaces where peers felt genuinely heard and understood. In these spaces, difficult conversations could unfold without fear of stigma, allowing peers to openly share their personal struggles and aspirations.

> *“They [fellows] were thinking of us and our health, remembering us… they care about us a lot.”* – peer, female

Supervisors, acknowledging the depth of these relationships, emphasized the importance of providing consistent mentorship and capacity-building support. They highlight how, once supported, fellows become role models themselves, leading by example and using their own experiences to guide others.

### Peer influence

Shared experiences between fellows and peers fostered a sense of belonging and emotional support. Peers described that the understanding derived from similar life journeys created an environment where they felt seen, heard, and inspired, and accepted them as positive role models who mirrored their challenges.

> *“We feel less stressed when we speak to the HIV positive person and share our problems with them and we also feel more comfortable with them.”* – peer, male

As a result of this connection, peers felt more receptive to encouragement and advice on healthy behaviors, such as adhering to their medication and staying physically active.

Supervisors reinforced this by noting that fellows’ lived experiences with HIV enabled them to form more profound and meaningful relationships with their peers than staff members. While these close connections provided essential support, they also burdened fellows emotionally. The empathy that made them effective mentors also made them vulnerable to emotional exhaustion, as fellows often found themselves reliving their hardships through the experiences of their peers.

> *“She’s [peer] the only daughter, and her mother died at a very young age. I don’t have any solution because my family has the same problem. I don’t have property. I too don’t have parents. So, I just started crying with her.”* – fellow, female

### Personal growth

Fellows indicated that personal growth is a dynamic process. Fellows described how their motivation developed through their internal belief in the ability to empower others, specifically using lived experiences to inspire others navigating similar challenges.

> *“I also wanted to show other children also they can do something…”* – fellow, female

Those who initially struggled with self-doubt, found strength in their relationship with others, cultivating resilience, specifically relational resilience, by leaning on the support of their peer network. Supervisors witnessed this transformation firsthand, observing fellows evolve from uncertain participants into confident leaders. They not only observed fellows acquiring practical skills such as time management, effective communication but also a deeper understanding of themselves and their community. Peers, too, perceived this potential for growth in the fellow role, seeing it as an opportunity for empowerment, education, and self-improvement. Those who once received support were eager to step into guiding roles themselves, reflecting an aspirational cycle of mentorship.

### Complex challenges

Fellows often grappled with financial instability and limited access to educational opportunities, which led them to highlight the need for career guidance and exposure to life beyond the program. Like the fellows, peers expressed uncertainty about their future, particularly transitioning out of the structured environments of CCIs and the fellowship.

> *“After finishing 12th, we are not sure if we will stay in [CCI] or not, and if they suddenly send us out and we cannot find a job, we will be alone. We are not aware of what support we will be getting.”* – peer, male

This uncertainty is exacerbated by the fact that, as Fellows observed, their peers face numerous challenges that could jeopardize a successful transition into adulthood.

> *“In this area, child marriage they’ll do. Like seven years they’ll do, then girls they won’t go school. Boys also, in second standard, they’ll say, ‘I don’t want to go to school,’ then their family members won’t allow them to go, and they’ll just sit there. Education problems are there, marriage problems, and financial problems.”* – fellow, female

Supervisors also recognized these concerns, identifying transition planning gaps, particularly job placement and long-term mentorship. They suggested that establishing a system for post-fellowship support, such as regular check-ins, networking opportunities, and job placement assistance, would help fellows successfully transition to life after the fellowship and maintain the connections they built during the program

Additionally, although fellows did develop practical skills throughout the fellowship, they still, at times, felt underprepared to fulfill their responsibilities, especially in the early stages. They described initial challenges with facilitating peer support, leading sessions, and completing administrative tasks such as writing reports, often struggling to find confidence in these new roles.

Supervisors also observed these difficulties and recommended setting clearer expectations and providing more structured preparation for fellows’ roles during the training process.

### Sustainability

Challenges related to financial instability and limited access to educational opportunities led fellows to highlight the need for career guidance and exposure to life beyond the program. Like the fellows, peers expressed uncertainty about their future, particularly transitioning out of structured environments of CCIs and the fellowship.

> *“Now we are in [CCI]. Here we get support but after we go out from [CCI] where will we get support from?”* – peer, male

Supervisors echoed this concern, identifying transition planning gaps, particularly career placement and long-term mentorship. They suggested that establishing a system for post-fellowship support, such as regular check-ins, networking opportunities, and job placement assistance, would help fellows successfully transition to life after the fellowship and maintain the connections they built during the program. Continuing this cycle of support, peers generally wanted to stay connected with the program and become future fellows. However, some had reservations about taking on a fellowship role due to concerns about their capabilities or the potential stigma associated with disclosing their HIV status.

> *“Fellows talk to everyone and share and I don’t freely share with others and that’s why I can’t work like them…Sometimes I want to but on the other side I feel nervous.”* – peer, female

## Discussion

The key findings from this qualitative study highlight how the I’mPossible Fellowship fosters personal growth, validation, and self-confidence among APHIV. (Figure 2). The fellowship, reinforced by peer support, ensures that services are accessible and responsive to the unique needs of APHIV. Our findings suggest that this model of peer support can lead to improved health behaviors, reduced stigma, better coping mechanisms, and a smoother transition into adulthood, resulting in sustained community impact (Figure 2). This holistic, multidimensional approach underscores the value of integrating peer mentorship into DSD frameworks to enhance long-term health outcomes and social well-being through two unique features: (1) service delivery that is more inclusive of other non-health-related domains and (2) focusing on the well-being of the individuals providing support.

**Figure 2.**
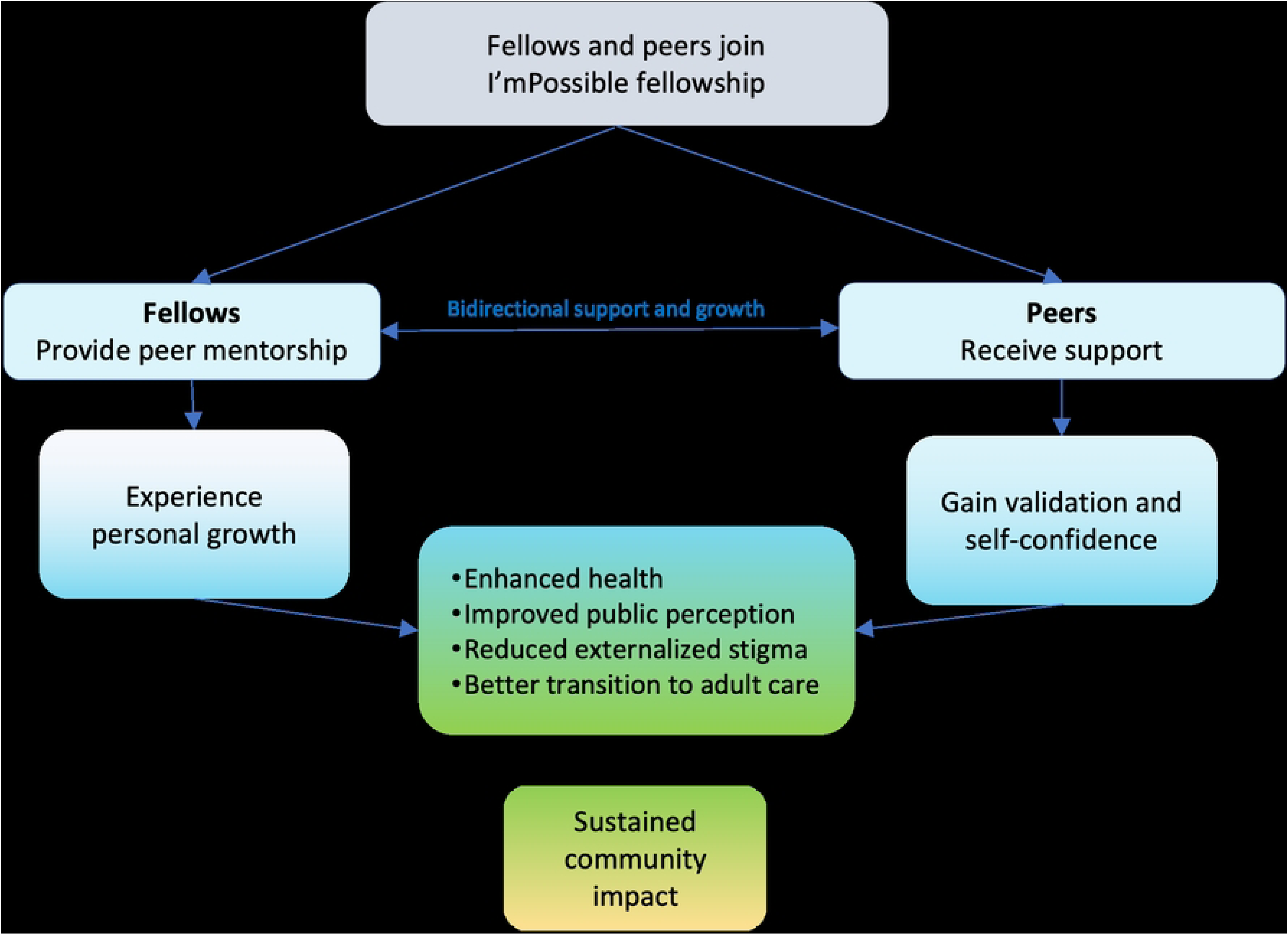
The journey of fellows and peers throughout the I’mPossible fellowship: insights from the study findings.

The I’mPossible fellowship, while aligned with known DSD frameworks, distinguishes itself through integrating education, vocational training, and community-based support into the peer mentorship framework, an evolution into what could be considered an advanced DSD approach (29). Earlier DSD models were designed to decentralize HIV services, promote task shifting, and enhance accessibility to medical treatment. Our approach, however, is expanding the definition of DSD to be more multilevel, person-centered, and community-driven, recognizing that sustained engagement in care requires interventions beyond just medical treatment (29,30). This next-generation DSD model acknowledges that social determinants of health, including poverty, stigma, mental health, and educational disparities, directly impact treatment adherence and overall well-being (30,31). As global HIV response efforts evolve, implementing holistic, youth-centered DSD models, like the I’mPossible fellowship, will be critical to ensuring that every YLHIV has the tools, networks, and opportunities to thrive.

A key feature of the I’mPossible fellowship is its emphasis on the well-being and sustainability of the peer mentors themselves. Unlike traditional models focusing solely on mentee outcomes, the fellowship recognizes that effective peer mentorship requires “caring for the carer” strategies (22). Fellows, many of whom have faced their own challenges with stigma, mental health, and treatment adherence, regardless of their leadership abilities, require sustained emotional support from supervisors and access to professional development opportunities to prevent emotional exhaustion. This approach strengthens the overall effectiveness of peer-led interventions and ensures that mentors can model resilience and self-efficacy for their peers.

Mentorship is the foundation of the fellowship and is driven by fellows’ sharing of lived experiences to provide emotional support and practical guidance to their peers. The bond that emerges, as a result, fosters a sense of psychological safety, which enables peers to discuss their challenges without fear of judgment and fellows to feel empowered to share their stories. This relational depth of mentorship emerged organically, reinforcing the essential components of peer support: informational, emotional, and affirmational support (28). Fellows provide informational support by educating peers on HIV management strategies, emotional support by offering empathy and encouragement, and affirmational support by validating traumatic and emotional experiences.

However, these bidirectional benefits, coupled with complex, systemic barriers, can hinder the sustainability and scalability of peer mentorship in HIV care. Stigma and discrimination, for example, deterred many APHIVs from accessing care. While fellows often acted as intermediaries between youth and healthcare services, fears of being discriminated against persisted in schools, workplaces, and residential living. Many APHIV anticipated facing limited education and employment opportunities because of prevailing community attitudes towards HIV. Evidence from existing literature suggests that this vulnerability can compel APHIV to prioritize immediate needs, such as financial stability over adherence and retention to care, threatening their long-term health management (31). These transition challenges can be addressed by integrating educational and vocational support to ensure peer mentorship translates into long-term sustainable health and social outcomes.

These findings align with global peer mentorship models that have successfully leveraged the power of trust and relatability of peers to improve health outcomes among YLHIV, although there are limited examples. Zimbabwe’s Zvandiri program has effectively integrated community adolescent treatment supporters (CATS) into healthcare settings, providing psychosocial support, health education, and care linkage services (19,22). Similarly, Zambia’s Project YES! saw measurable improvements in viral suppression and mental health outcomes after placing peer mentors in HIV clinics (21,32). These programs reinforce that peer-led approaches within DSD models support positive treatment outcomes, adherence, and well-being for YLHIV in low- and middle-income settings.

While our study offers valuable insights into youth-led interventions for HIV care, we acknowledge some limitations. Our research focused on the first batch of fellows within the initial 18 months of implementing the DSD model. As this was an early phase with small numbers conducted when the model was still being refined, the experiences captured reflect preliminary lessons. The study is highly contextual, and findings may vary in different healthcare infrastructures or stages of DSD model implementation. Nevertheless, our study offers essential strengths, including its youth-centered participatory approach and the rich qualitative data drawn from the direct experiences of both mentors and mentees. These insights provide valuable guidance for refining peer-led mentorship models and tailoring DSD approaches to meet the evolving needs of YLHIV better.

In conclusion, the I’mPossible fellowship represents a significant advancement of community-based DSD models through the operationalization of structured peer mentorship. By integrating informational, emotional, and affirmational support with critical psychosocial, educational, and social dimensions, the program demonstrates how DSD can be expanded to best address the needs of YLHIV. Future research should focus on scalability, sustainability, and long-term impact, particularly in addressing economic barriers and stigma-related challenges that hinder optimal youth transition into adult care and successful integration into general society.

## Data Availability

The data analyzed in this study were collected as part of a qualitative research project. Relevant excerpts from participant transcripts are included within the paper. De-identified data used in this study may be accessed upon reasonable request to the corresponding author.

## Acknowledgments

The authors acknowledge the RISHI Foundation and Sneha Charitable Trust, as well as the participating childcare institutions and participants, for their contributions to this research. The authors wish to recognize travel support from the Johns Hopkins Center for Global Health and the Girish and Rishi Himangi Scholarship through the Gupta-Klinsky India Institute at Johns Hopkins University. The study was partly funded through the Rishi Children’s Fund at the Department of International Health at Johns Hopkins Bloomberg School of Public Health and support received by LG from the National Institute on Drug Abuse of the National Institutes of Health (K23DA057151).

## Abbreviations

APHIV: Adolescents and Young Adults with Perinatally Acquired HIV
ART: Antiretroviral Treatment
CCI: Childcare Institution
DSD: Differentiated Service Delivery
FGD: Focus Group Discussion
HIV: Human Immunodeficiency Virus
IDI: In-Depth Interview
SEM: Socio-ecological Model
YLHIV: Youth Living with HIV

## References

1. UNICEF, UNAIDS. Adolescent HIV treatment. 2024 [cited 2024 Jun 28]. Available from: https://data.unicef.org/topic/hivaids/adolescent-hiv-treatment/

2. Slogrove AL, Sohn AH. The global epidemiology of adolescents living with HIV: time for more granular data to improve adolescent health outcomes. Curr Opin HIV AIDS. 2018 May 1;13(3):170–8.

3. Sohn AH, Davies M. Adults with perinatally acquired HIV in low- and middle-income settings: time for a generational shift in HIV care and global guidance. J Int AIDS Soc. 2024 Jul 22;27(7).

4. Mellins CA, Malee KM. Understanding the mental health of youth living with perinatal HIV infection: Lessons learned and current challenges. J Int AIDS Soc. 2013;16.

5. Bhana A, Kreniske P, Pather A, Abas MA, Mellins CA. Interventions to address the mental health of adolescents and young adults living with or affected by HIV: state of the evidence. J Int AIDS Soc. 2021 Jun 1;24(Suppl 2). Available from: /pmc/articles/PMC8222850/

6. Remien RH, Stirratt MJ, Nguyen N, Robbins RN, Pala AN, Mellins CA. Mental health and HIV/AIDS. AIDS. 2019 Jul 15;33(9):1411–20.

7. Vreeman RC, McCoy BM, Lee S. Mental health challenges among adolescents living with HIV. J Int AIDS Soc. 2017 May 16;20(Suppl 3).

8. Chadda RK. Youth & mental health: Challenges ahead. Indian J Med Res. 2018 Oct 1;148(4):359–61.

9. Haas AD, Technau KG, Pahad S, Braithwaite K, Madzivhandila M, Sorour G, et al. Mental health, substance use and viral suppression in adolescents receiving ART at a paediatric HIV clinic in South Africa. J Int AIDS Soc. 2020 Dec 1;23(12).

10. Sudjaritruk T, Aurpibul L, Songtaweesin WN, Narkpongphun A, Thisayakorn P, Chotecharoentanan T, et al. Integration of mental health services into HIV healthcare facilities among Thai adolescents and young adults living with HIV. J Int AIDS Soc. 2021 Feb 1;24(2).

11. Dow DE, Turner EL, Shayo AM, Mmbaga B, Cunningham CK, O’Donnell K. Evaluating mental health difficulties and associated outcomes among HIV-positive adolescents in Tanzania. AIDS Care. 2016 Jul 2;28(7):825–33.

12. Mavhu W, Willis N, Mufuka J, Bernays S, Tshuma M, Mangenah C, et al. Effect of a differentiated service delivery model on virological failure in adolescents with HIV in Zimbabwe (Zvandiri): a cluster-randomised controlled trial. Lancet Glob Health. 2020 Feb;8(2):e264–75.

13. Mackenzie RK, Van Lettow M, Gondwe C, Nyirongo J, Singano V, Banda V, et al. Greater retention in care among adolescents on antiretroviral treatment accessing “Teen Club” an adolescent-centred differentiated care model compared with standard of care: a nested case-control study at a tertiary referral hospital in Malawi. 2017. 20(3):e25028.

14. World Health Organization. Adolescent friendly health services for adolescents living with HIV: from theory to practice. Geneva: World Health Organization; 2019 [cited 2024 Jun 28]. (WHO/CDS/HIV/19.39). Available from: https://www.who.int/publications/i/item/adolescent-friendly-health-services-for-adolescents-living-with-hiv

15. Bernays S, Tshuma M, Willis N, Mvududu K, Chikeya A, Mufuka J, et al. Scaling up peer-led community-based differentiated support for adolescents living with HIV: keeping the needs of youth peer supporters in mind to sustain success. 2020. 23 Suppl 5(Suppl 5):e25570.

16. Barker D, Enimil A, Galárraga O, Bosomtwe D, Mensah N, Thamotharan S, Henebeng E, Brown L, Kwara A. In-Clinic Adolescent Peer Group Support for Engagement in Sub-Saharan Africa: A Feasibility and Acceptability Trial. J Int Assoc Provid AIDS Care. 2019 Jan-Dec;18. doi: 10.1177/2325958219835786.

17. Genberg BL, Shangani S, Sabatino K, Rachlis B, Wachira J, Braitstein P, et al. Improving Engagement in the HIV Care Cascade: A Systematic Review of Interventions Involving People Living with HIV/AIDS as Peers. AIDS Behav. 2016 Oct 1;20(10):2452–63.

18. Miyingo C, Mpayenda T, Nyole R, Ayinembabazi J, Ssepuuya M, Ssebuwufu EM, et al. HIV Treatment and Care of Adolescents: Perspectives of Adolescents on Community-Based Models in Northern Uganda. HIV/AIDS - Res Palliative Care. 2023 Mar;15:105–14.

19. Willis N, Napei T, Armstrong A, Jackson H, Apollo T, Mushavi A, et al. Zvandiri—Bringing a Differentiated Service Delivery Program to Scale for Children, Adolescents, and Young People in Zimbabwe. JAIDS. 2018 Aug 15;78(2):S115–23.

20. Rencken CA, Harrison AD, Mtukushe B, Bergam S, Pather A, Sher R, et al. “Those People Motivate and Inspire Me to Take My Treatment.” Peer Support for Adolescents Living With HIV in Cape Town, South Africa. J Int Assoc AIDS Care. 2021 Jan 1;20. doi: 10.1177/23259582211000525.

21. Denison JA, Burke VM, Miti S, Nonyane BAS, Frimpong C, Merrill KG, et al. Project YES! Youth Engaging for Success: A randomized controlled trial assessing the impact of a clinic-based peer mentoring program on viral suppression, adherence and internalized stigma among HIV-positive youth (15-24 years) in Ndola, Zambia. PLoS One. 2020 Apr;15(4).

22. Tailor LS, Angell J, Hasan S, Low S, Willis N, Mutsinze A, Chitiyo V, Kuchocha P, Logie CH. Bolstering Access to HIV-Related Health care in Zimbabwe Among Young Mothers Living With HIV: Lessons Learned on HIV Health Promotion From Zvandiri’s Young Mentor Mother Program. Health Promot Pract. 2024 Sep 30. doi: 10.1177/15248399241278974.

23. UNICEF. HIV Statistics - Global and Regional Trends. 2024. Available from: https://data.unicef.org/topic/hivaids/global-regional-trends/

24. National AIDS Control Organisation. Sankalak: Status of National AIDS & STD Response (Fifth edition, 2023). New Delhi; 2023 [cited 2024 Oct 17]. Available from: https://naco.gov.in/sites/default/files/Sankalak_Booklet_Fifth_Edition_2023.pdf

25. Columbia University. Adolescent HIV Care and Treatment: A Training Curriculum for Health Workers. 2012 [cited 2025 Feb 24]. Available from: https://icap.columbia.edu/tools_resources/adolescent-hiv-care-and-treatment-a-training-curriculum-for-health-workers/

26. Bronfenbrenner U. The ecology of human development: Experiments by nature and design. Harvard University Press; 1979.

27. Braun V, Clarke V. Using thematic analysis in psychology. Qual Res Psychol. 2006 Jan;3(2):77–101.

28. Vogl S, Schmidt EM, Zartler U. Triangulating perspectives: ontology and epistemology in the analysis of qualitative multiple perspective interviews. Int J Soc Res Methodol. 2019 Nov 2;22(6):611–24.

29. Ehrenkranz P, Grimsrud A, Holmes CB, Preko P, Rabkin M. Expanding the Vision for Differentiated Service Delivery: A Call for More Inclusive and Truly Patient-Centered Care for People Living With HIV. J Acquir Immune Defic Syndr. 2021 Feb 1;86(2):147–52.

30. Menza TW, Hixson LK, Lipira L, Drach L. Social Determinants of Health and Care Outcomes Among People With HIV in the United States. Open Forum Infect Dis. 2021 Jun 22;8(7):ofab330.

31. Holtzman CW, Brady KA, Yehia BR. Retention in Care and Medication Adherence: Current Challenges to Antiretroviral Therapy Success. Drugs. 2015 Apr 20;75(5):445–54.

32. Burke VM, Frimpong C, Miti S, Mwansa JK, Abrams EA, Merrill KG, et al. “It must start with me, so it started with me”: A qualitative study of Project YES! youth peer mentor implementing experiences supporting adolescents and young adults living with HIV in Ndola, Zambia. PLoS One. 2022 Feb 1;17(2):e0262087.

